# Budget Impact of Replacing In-Laboratory Polysomnography With Comprehensive Home Polysomnography Using the Onera Sleep Test System in a U.S. Commercial Health Plan

**DOI:** 10.64898/2026.05.13.26352915

**Authors:** Jennifer Hinkel, Shivani Modi, Arka Ray, Joel Brill

## Abstract

**Background:** In-laboratory polysomnography (PSG) remains the diagnostic reference standard for sleep disorders but is resource-intensive and capacity-constrained. Limited-channel home sleep apnea testing (HSAT) improves access and reduces costs compared to in-laboratory polysomnography, but underestimates disease severity due to its inability to measure true sleep time and cannot identify non-respiratory sleep disorders including periodic limb movement disorder and parasomnias.^1–5^ Comprehensive home polysomnography (hPSG) may preserve diagnostic fidelity while reducing system costs, improving access for patients unable to attend laboratory-based studies, and shortening time to diagnosis and therapy initiation.

**Objective:** To estimate the short-term budget impact to a U.S. commercial health plan of substituting an appropriately selected proportion of in-laboratory PSG with comprehensive hPSG using the Onera Sleep Test System (STS).

**Methods:** We developed a transparent budget impact model following ISPOR good practice guidelines for a hypothetical 1-million-member commercial plan. The model estimates the annual diagnostic population (top-of-funnel) using age- and sex-stratified prevalence, an undiagnosed fraction of 85%, symptom prevalence among undiagnosed individuals (30%), and an annual testing rate (12%).^2–3^ Baseline costs reflect current diagnostic pathways using HSAT (50% first-line) and in-laboratory PSG (50% first-line), including HSAT-to-PSG escalations (20%) and PSG repeats (4%). The intervention scenario substitutes a defined share of in-laboratory PSG and selected HSAT with Onera hPSG. Scenario and sensitivity analyses explore parameter uncertainty.

**Results:** In the base case, approximately 4,364 individuals entered the OSA diagnostic workflow annually. Baseline diagnostic costs were estimated at $6.23 PMPM, comprising $5.45 million in PSG costs and $0.79 million in HSAT costs. Introducing Onera hPSG (30% PSG replacement, 5% HSAT replacement in Year 1) reduced per member costs to $5.66 PMPM, yielding net savings of $0.57 PMPM ($567,262 annually). In Year 3 scenarios (60% PSG, 10% HSAT replacement), savings increased to $1.64 PMPM (approximately $1.64 million annually). Sensitivity analyses demonstrated net savings ranging from $0.03 to $8.05 PMPM, depending on adoption levels.

**Conclusions:** Partial substitution of in-laboratory PSG with Onera hPSG may yield incremental budget savings for U.S. commercial payers while maintaining access to full polysomnographic assessment. Results support further payer-specific analyses incorporating real-world utilization and downstream outcomes.

## Introduction

Obstructive sleep apnea (OSA) and related sleep disorders represent a large, underdiagnosed source of morbidity and healthcare utilization in the United States. Global epidemiologic analyses suggest that nearly one billion adults worldwide have OSA, with U.S. prevalence estimates ranging from 9% to 38% depending on age, sex, and diagnostic thresholds.^1,2^ In contrast, large U.S. administrative claims analyses show diagnosed prevalence in commercially insured populations of approximately 3.4%, underscoring a substantial diagnosis gap.^3^

This divergence between epidemiologic and claims-based prevalence reflects structural constraints in diagnostic capacity rather than low disease incidence. Population-based studies from the Wisconsin Sleep Cohort and Sleep Heart Health Study estimated that up to 80% of individuals with moderate-to-severe OSA remained undiagnosed despite adequate access to healthcare.^4,5^ More recent analyses suggest this figure can reach 85% when accounting for evolving prevalence estimates and diagnostic patterns.^6^ In-laboratory polysomnography (PSG), while considered the reference standard, is constrained by facility capacity, workforce shortages, and high per-study costs, resulting in prolonged wait times and diagnostic attrition.^7^

Home sleep apnea testing (HSAT) has expanded access by shifting limited-channel diagnostics into the home. Randomized trials and economic evaluations have shown HSAT-based pathways to be non-inferior to PSG for uncomplicated, high-pretest-probability OSA in selected patients.^7^ However, although some HSAT devices include methods to estimate sleep stages, EEG remains the direct measurement and “gold standard” for sleep staging and assessment of overall sleep architecture. As a result, HSAT can underestimate disease severity, fail to diagnose non-respiratory sleep disorders such as periodic limb movement disorder (PLMD), parasomnias, and comorbid insomnia and sleep apnea (COMISA), and result in frequent escalation to PSG after negative or indeterminate results.^8^ These limitations dilute both clinical and economic advantages at the system level.

Recent advances in patch-based sensing technology enable unattended, comprehensive hPSG that captures the full signal set required for sleep staging and respiratory event detection. Onera hPSG systems aim to combine the diagnostic completeness of PSG with the accessibility of home testing. A recent multicenter validation study demonstrated strong agreement between Onera hPSG and in-laboratory PSG across sleep architecture and apnea-hypopnea metrics.^9^ Systematic reviews have examined the technical feasibility of hPSG more broadly.^10^

For U.S. payers, however, adoption decisions hinge less on technical validity and more on budget impact. Budget impact analyses (BIA) complement cost-effectiveness studies by answering a pragmatic question: what happens to plan spending if a new technology is introduced into existing care pathways? The objective of this study is to present a transparent, adaptable budget impact modeling framework that payers can parameterize to their own covered populations. Rather than asserting a single universal savings estimate, the model emphasizes diagnostic flow, escalation, and substitution effects to support payer decision-making regarding comprehensive hPSG adoption.

## Methods

### Analytic Framework

This analysis follows ISPOR good practice guidance for budget impact analysis, emphasizing transparency and short-term payer relevance.^11^ The model is modular by design to support payer-specific adaptation. The Excel-based model is available as supplementary material.

### Perspective and Time Horizon

The perspective is that of a U.S. commercial health plan. The time horizon spans one to five years, consistent with annual budgeting cycles and standard BIA conventions. Only direct diagnostic testing costs are included; downstream treatment and outcome effects were not modeled.

### Target Population and Diagnostic Funnel

We modeled a hypothetical commercial plan with 1,000,000 covered lives, distributed by age and sex consistent with typical commercial demographics (Table 1). The annual diagnostic population represents members newly entering evaluation for suspected OSA. This population is derived using age- and sex-stratified true prevalence estimates, an undiagnosed fraction, symptom prevalence among undiagnosed individuals, and an annual testing rate reflecting real-world care-seeking behavior.

**Table 1.** Top-of-Funnel: Annual OSA Diagnostic Population by Age and Sex.

| Age-Sex Band | Annual Top-of-Funnel (N) |
| --- | --- |
| Male 18-34 | 704 |
| Female 18-34 | 404 |
| Male 35-49 | 1,102 |
| Female 35-49 | 624 |
| Male 50-64 | 918 |
| Female 50-64 | 612 |
| **Total** | **4,364** |

True prevalence estimates were selected from peer-reviewed epidemiologic studies using apnea-hypopnea index (AHI)-based definitions. We applied age- and sex-specific prevalence consistent with established gradients: 10% for males 18-34, 6% for females 18-34, 20% for males 35-49, 12% for females 35-49, 30% for males 50-64, and 20% for females 50-64.^2^

The model assumes that 85% (range: 68-91%) of true OSA cases remain undiagnosed, consistent with reported data showing that diagnosed prevalence (3.4%) substantially underestimates true prevalence.^3–6^ Among these undiagnosed individuals, approximately 30% (range: 25-35%) are symptomatic, representing the subset likely to consider or seek testing. Approximately one-third of symptomatic undiagnosed individuals are expected to reach testing within three years, yielding an annual testing rate of 12% (range: 8-18%).

### Baseline Diagnostic Pathway

In the baseline scenario, patients entering the diagnostic pathway initiate testing with either HSAT or in-laboratory PSG. 50% were assumed to start with HSAT and 50% with PSG, consistent with AASM 2017 guidelines specifying HSAT as appropriate first-line in uncomplicated, high-probability OSA, with PSG for complex cases.^7^ HSAT studies may escalate to PSG when results are negative, indeterminate, or technically inadequate in the setting of persistent clinical suspicion. An HSAT-to-PSG escalation rate of 20% was applied based on guideline-consistent practice.^7,8^ A proportion (4%) of PSGs were assumed to repeat due to insufficient sleep or technical failure.

### Intervention Scenario: Introduction of hPSG

The intervention scenario introduces Onera hPSG as a substitute for a proportion of in-laboratory PSGs and a limited share of HSATs. In Year 1, 30% of in-laboratory PSG studies (both PSG-first and PSG follow-ups from HSAT escalation) and 5% of HSATs were replaced by hPSG. Year 3 scenarios assumed 60% PSG and 10% HSAT replacement. Residual escalation to in-laboratory PSG (2%) and repeat rates (2%) were applied conservatively to hPSG, reflecting fewer technical failures due to patient self-application and multi-night capability.

### Unit Costs

Unit costs were derived from Medicare fee schedules and commercial payer data where available. Base case costs were: HSAT $360 per study (CPT 95806/HCPCS G0398; sensitivity range $200-$500), in-laboratory PSG $2,000 per study (CPT 95810; sensitivity range $1,200-$3,000), and hPSG $1,400 per study (sensitivity range $1,200-$1,600). Costs include technical and professional components. The hPSG cost represents a commercial midpoint between HSAT and in-laboratory PSG. Table 2 presents the full parameter set.

**Table 2.** Model Parameters and Sources.

| Parameter | Base Case | Range | Source |
| --- | --- | --- | --- |
| Plan population | 1,000,000 | Plan-specific | HEOR convention |
| Undiagnosed share | 85% | 68-91% | Young 2002; Senaratna 2017 |
| Symptomatic share | 30% | 25-35% | Assumption |
| Annual testing rate | 12% | 8-18% | Derived assumption |
| HSAT-first share | 50% | 30-70% | Kapur 2017 (AASM) |
| HSAT-to-PSG escalation | 20% | 10-30% | Kapur 2017; Bianchi 2017 |
| PSG repeat rate | 4% | 4-6% | Assumption |
| hPSG replaces PSG (Year 1) | 30% | 10-70% | Scenario assumption |
| hPSG replaces HSAT (Year 1) | 5% | 0-25% | Scenario assumption |
| hPSG repeat rate | 2% | 1-3% | Assumption (multi-night) |
| HSAT unit cost | \$360 | \$200-\$500 | Commercial midpoint |
| In-laboratory PSG unit cost | \$2,000 | \$1,200-\$3,000 | Kim 2015; commercial data |
| hPSG (Onera) unit cost | \$1,400 | \$1,200-\$1,600 | Manufacturer estimate |

*HSAT = home sleep apnea testing; PSG = polysomnography; hPSG = home polysomnography*

**Table 3.** Budget Impact Results: Baseline vs. Onera Scenarios.

| Component | Baseline | Onera Year 1 | Onera Year 3 |
| --- | --- | --- | --- |
| Total PSG costs | \$5,445,723 | \$3,665,390 | \$1,396,339 |
| Total HSAT costs | \$785,441 | \$746,169 | \$691,188 |
| Onera hPSG costs | \$0 | \$1,252,342 | \$2,504,683 |
| **Total diagnostic costs** | **\$6,231,164** | **\$5,663,901** | **\$4,592,210** |
| **PMPM** | **\$6.23** | **\$5.66** | **\$4.59** |
| **PMPM savings vs. baseline** | **—** | **\$0.57** | **\$1.64** |

### Outcomes

Primary outcomes include total annual diagnostic costs and per-member-per-month (PMPM) impact. Secondary outcomes include modality-specific cost shifts and test volumes.

### Sensitivity and Scenario Analyses

One-way sensitivity analyses varied each parameter individually across plausible ranges (Table 2). Scenario analyses examined Year 3 adoption (60% PSG substitution, 10% HSAT substitution) and extreme scenarios (up to 90% substitution). The model structure allows payers to input plan-specific values.

## Results

### Baseline Diagnostic Volumes and Costs

In the modeled 1-million-member plan, the annual diagnostic population for suspected OSA was 4,364 individuals. Under baseline assumptions, this generated 2,182 HSAT-first studies, 2,182 PSG-first studies, 436 HSAT-to-PSG escalations, and 105 PSG repeats. Baseline diagnostic spending for OSA was estimated at $6.23 million annually ($6.23 PMPM), with in-laboratory PSG accounting for $5.45 million (approximately 87%) and HSAT for $0.79 million (13%) of total costs.

### Budget Impact with Onera

Introduction of Onera hPSG at Year 1 adoption levels (30% PSG replacement, 5% HSAT replacement) reduces total diagnostic spending to approximately $5.66 million annually ($5.66 PMPM), representing net savings of $0.57 PMPM ($567,262 annually). Savings are primarily driven by a $1.78 million reduction in in-laboratory PSG costs, partially offset by $1.25 million in Onera hPSG testing costs, along with a slight decrease in HSAT spending.

In Year 3 scenarios assuming broader adoption (60% PSG replacement, 10% HSAT replacement), savings increase to approximately $1.64 PMPM ($1.64 million annually).

### Sensitivity Analyses

Results were most sensitive to the proportion of PSG replaced by hPSG and to the unit cost differential between PSG and hPSG. Across sensitivity ranges, per member savings ranged from near cost neutrality ($0.03 PMPM) to substantial savings ($8.05 PMPM) under high-substitution scenarios. Even under conservative assumptions, the intervention did not increase total plan spending.

## Discussion

This analysis demonstrates that substituting an appropriately selected proportion of in-laboratory PSG with Onera hPSG can generate incremental short-term savings for U.S. commercial payers. Importantly, the primary contribution of this work is not a single point estimate of savings, but the presentation of a transparent, flow-based budget impact model that can be adapted to payer-specific populations and utilization patterns.

The modeled savings are driven by replacing higher-cost laboratory studies ($2,000) with lower-cost home-based studies ($1,400) that deliver comparable diagnostic information. The $600 per-study differential, applied across approximately 30% of PSG volume in Year 1, generates the observed savings. Unlike HSAT-centric strategies, which trade diagnostic completeness for access, comprehensive hPSG preserves full polysomnographic assessment while reducing reliance on resource-intensive laboratory infrastructure.

Claims-based analyses demonstrate that current diagnostic pathways capture only a fraction of true disease burden, reflecting constrained diagnostic capacity rather than low prevalence.^3^ The model explicitly addresses payer concerns regarding induced utilization by holding total diagnostic demand constant and examining substitution effects within existing pathways. The top-of-funnel calculation (4,364 individuals annually in a 1M-member plan) reflects observed diagnostic patterns, not a target for expanded testing.

From a clinical perspective, the model assumes that comprehensive hPSG can provide diagnostic information comparable to in-laboratory PSG while expanding access and convenience. However, this assumption rests on limited real-world evidence. The primary validation study for the Onera hPSG demonstrated agreement with in-laboratory PSG, but was conducted in Germany using simultaneous recording rather than unsupervised home deployment.

Additionally, the internal manufacturer data for the Onera hPSG system (STS 1) indicate a patient study completion rate of 94% (Onera Health data on file).

### Limitations

Several limitations warrant consideration. First, results are based on modeled assumptions rather than plan-specific claims data and do not stratify by comorbidity, severity, or geography. Second, the clinical validation evidence for Onera hPSG derives primarily from a single industry-funded multicenter study conducted in Germany.^9^ Real-world performance in unsupervised U.S. home settings can differ. Third, downstream benefits of earlier diagnosis (e.g., reduced cardiovascular events, motor vehicle accidents, healthcare utilization) were not included, making estimates conservative from a societal perspective but appropriate for short-term payer budgeting. Fourth, Medicare populations were not modeled; budget impact may differ in elderly populations with different prevalence and comorbidity profiles. Fifth, the hPSG unit cost ($1,400) reflects manufacturer estimates rather than established reimbursement rates.

### Future Research

Future work should apply this modeling framework to payer-specific administrative claims data, enabling calibration of prevalence, testing rates, escalation probabilities, and unit costs to observed experience. Prospective U.S. studies of hPSG performance would strengthen clinical assumptions. Longer time horizons incorporating downstream treatment initiation, adherence, and outcomes would provide a more complete economic picture.

## Conclusions

Adopting a comprehensive hPSG with Onera in an appropriately selected subset of patients using in-laboratory PSG can yield incremental PMPM savings ($0.57 in Year 1, potentially $1.64 by Year 3) for U.S. commercial health plans while maintaining diagnostic completeness. The transparent, adaptable model structure allows payers to evaluate budget impact using their own census, utilization, and cost data.

## Data Availability

All data produced in the present study are available upon reasonable request to the authors

## Notes

**Conflict of Interest Disclosure** The authors declare no individual competing interests.

**Funding** This study was supported by Onera Health, Inc., Palo Alto, USA. The funder provided input on study design but had no role in data analysis, interpretation, or the decision to submit for publication.

### Competing Interest Statement

Sumis Partners, Inc. was contracted by Onera Health, Inc. to conduct this analysis. Authors Hinkel, Modi, and Ray are employees or contractors of Sigla Sciences, and author Brill is an employees of Predictive Health; both Sigla Sciences and Predictive Health are affiliates of Sumis Partners, Inc. Sumis Partners, Inc. and its affiliates provide consulting services to life sciences companies, including diagnostic and device firms. The authors have no other competing interests to declare.

### Funding Statement

This study was supported by Onera Health, Inc., Palo Alto, USA. The funder provided input on study design but had no role in data analysis, interpretation, or the decision to submit for publication.

